# Flattening the COVID 19 curve in susceptible forest indigenous tribes using SIR model

**DOI:** 10.1101/2020.05.22.20110254

**Authors:** Andrio Adwibowo

## Abstract

COVID 19 is a global threat and globally spreading. The international cooperation involving indigenous peoples and local communities is urgently required in joint prevention to control the epidemic. Currently, many indigenous populations are continuing to face COVID 19. This study is concerned about the dynamic of COVID 19 pandemic among indigenous populations living in the remote Amazon rainforest enclaves. Using the Susceptible Infectious Recovered (SIR) model, the spread of the COVID 19 under 3 intervention scenarios (low, moderate, high) is simulated and predicted in indigenous tribe populations. The SIR model forecasts that without intervention, the epidemic peak may reach within 10-20 days. Nonetheless the peak can be reduced with strict interventions. Under low intervention, the COVID 19 cases are reduced to 73% and 56% of the total populations. While, in the scenario of high intervention, the COVID 19 peaks can be reduced to values ranging from 53% to 15%. To conclude, the simulated interventions tested by SIR model have reduced the pandemic peak and flattened the COVID 19 curve in indigenous populations. Nonetheless, it is mandatory to strengthen all mitigation efforts, reduce exposures, and decrease transmission rate as possible for COVID 19 containment.

## 1. Introduction

One of potential threats to indigenous populations is spread of communicable diseases. It is theoritized that a colonization of the New World has responsibility for delivering diseases and lead to the catastrophic depopulation to indigenous populations (Crosby 1976, Joralemon 1982). During early colonization, many indigenous populations had no prior exposure to pathogens that had become common outside indigenous population. In such populations, epidemics caused by acute infectious disease can be extremely severe, resulting in high mortality, and complete social disruption. In particular small indigenous communities, this situation occurred not only at the time of initial colonization, yet it is still happened repeatedly until present time.

Cases of communicable diseases in indigenous populations have reportedly widely, from Africa (Ohenjo *et al*. 2006, Liddell *et al*. 2005), Australia, to South America. In Africa indigenous population of the central African pygmy peoples and the San of southern Africa, poor health status is widely recognized. In Australia continent, communicable diseases explained 30% of the health gap among Aboriginal and Torres Strait islander populations (Vos *et al*. 2009).

A most comprehensive study on vulnerability of indigenous populations towards communicable diseases can be found in study by Walker *et al*. (2015). Their studies reported 117 epidemics that affected 59 different indigenous societies living in Amazon rainforest and caused over 11000 deaths between 1875 and 2008. Those epidemics including measles, influenza, and malaria with proportions were 37%, 25%, and 13%. The others epidemics that were below 10% included tuberculosis (7%), hepatitis (6%), smallpox (5%), chicken pox, pertussis, polio, and cholera.

The Amazon indigenous population related COVID 19 study is still limited and only a few literature available (Baumgartner *et al*. 2020, Ortiz-Prado *et al*. 2020). Eventhough the study is still limited, nonetheless the available literatures have provided the magnitude of COVID 19 pandemic among indigenous populations. For that reasons, this study aims to investigate the dynamic of COVID 19 pandemic among indigenous populations living in the remote Amazon rainforest enclaves. The investigations are made using the Susceptible Infectious Recovered (SIR) model combined with several intervention scenarios.

## 2. Methodology

### 2.1. Data collection

The study populations were the indigenous tribes living in the rainforest enclaves along the Amazon river (Figure 1). Those enclaves included Lagartococha, Yasuni, and Callarú. The data collected from the indigenous tribes were total population and COVID 19 confirmed cases. The data were obtained from the published data provided by Indigenous People Articulation, Regional Organization of Indigenous Peoples of the East, and The National Institute of Statistics and Informatics (INEI) websites.

**Figure 1.**
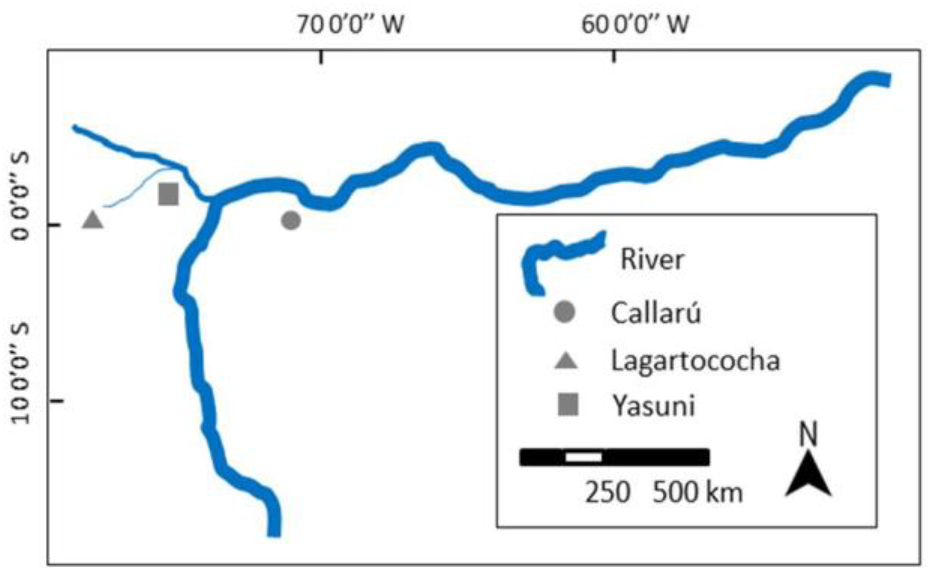
The geographical locations of indigenous tribes living in the rainforest enclaves along the Amazon river.

### 2.2. SIR model

In this study, a well known Susceptible Infectious Recovered (SIR) model was adopted (Jo *et al*. 2020, Zhao *et al*. 2020). In dealing with the current COVID 19 pandemic, SIR is considered as a versatile compartmental mathematical tool to model any pandemic dynamic. The model helps us to understand kinetics of pandemic with any specific aspects and the SIR model is also simple to understand and has clear interpretations (Wagh *et al*. 2020). In the standard SIR model (Waqas *et al*. 2020), the total population (N) is divided into three compartments. The Susceptible (S), the fraction of the total population that is vulnerable and at a risk of being infected. The Infected (I), the population who has been infected. The Recovered (R), the fraction of the total population that recovers. In the SIR model, there are several input parameters need to be considered. Those parameters denote as β and γ. According to Belfin *et al*. (2020), β is the effective transmission rate and γ is the removal rate (recovery). According to Mahmud and Lim (2020), γ is denoted as 1/d with d is the duration of recovery in day. The specific description of SIR model is as follows:

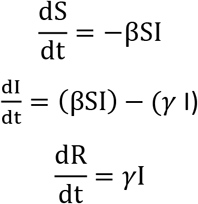

### 2.3. Intervention scenarios

The scenario analysis was used to observe the shape of COVID 19 SIR curve based on several simulated interventions applied. Likewise, simulations were conducted based on the scaled epidemic prevention and control intervention intensities ranging from low (25%), moderate (50%), and high (75%).

## 3. Results

The indigenous tribe populations and COVID 19 cases in Amazon rainforest enclaves including Lagartococha, Callarú, and Yasuni are presented in the Figure 2. The highest tribe population was observed in the Callarú and Lagartococha has the lowest population. Regarding the COVID 19 cases, the Callarú population has the highest cases in comparison to other populations. Until May 12, 2020, there were 50 cases reported. The second large COVID 19 cases among tribe population were observed in Lagartococha enclaves. Since May 5, 2020, there were already 15 cases in Lagartochocha population. Yasuni enclave has the lowest cases. Among Yasuni’s population equal to 2000 people, the first cases was reported on the last May 4, 2020.

**Figure 2.**
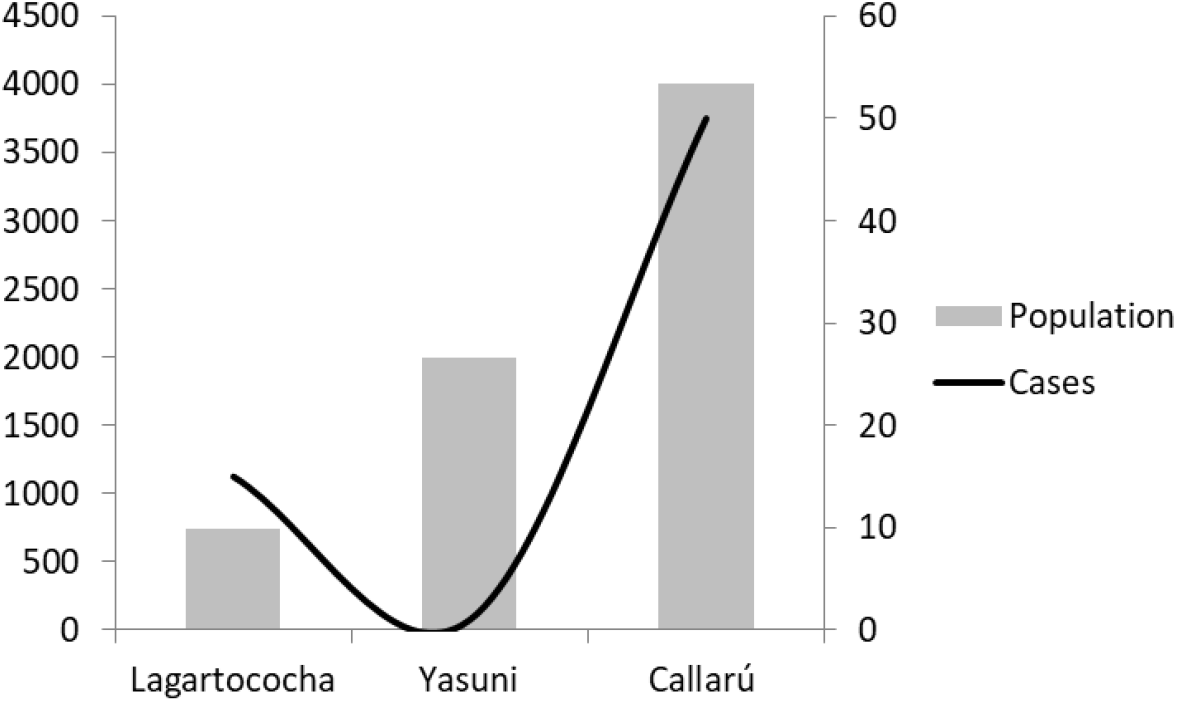
The indigenous tribe populations (0-4500) and COVID 19 cases (0-60) in Amazon rainforest.

The Figure 3 shows the SIR model applied to forecast the COVID 19 cases in Lagartococha and Yasuni rainforest enclaves based on the available data. From Figure 3, it is clear that the COVID 19 cases will grow rapidly over time resulting a vastly infected population both in Lagartococha and Yasuni. For the small tribe population as observed in Lagartococha, the COVID 19 is estimated will reach it peaks in less than 10 days. After that the infection rates will decrease and and the recovery rates will increase however at slow rates. The similar patters are also observed in the larger population as can be observed in the SIR model available for Yasuni population. This tribe population is estimated will also experience high increase of COVID 19 cases between 10 and 20 days which is more longer than what happens to small tribe population in this case the Lagartococha. Based on the SIR model, it is estimated that during its peaks, the highest cases for the Lagartococha population are between 86% to 93% of the total population equal to 744 people. While for the Yasuni population, the highest cases are equal to 66% of its total population (2000 people).

**Figure 3.**
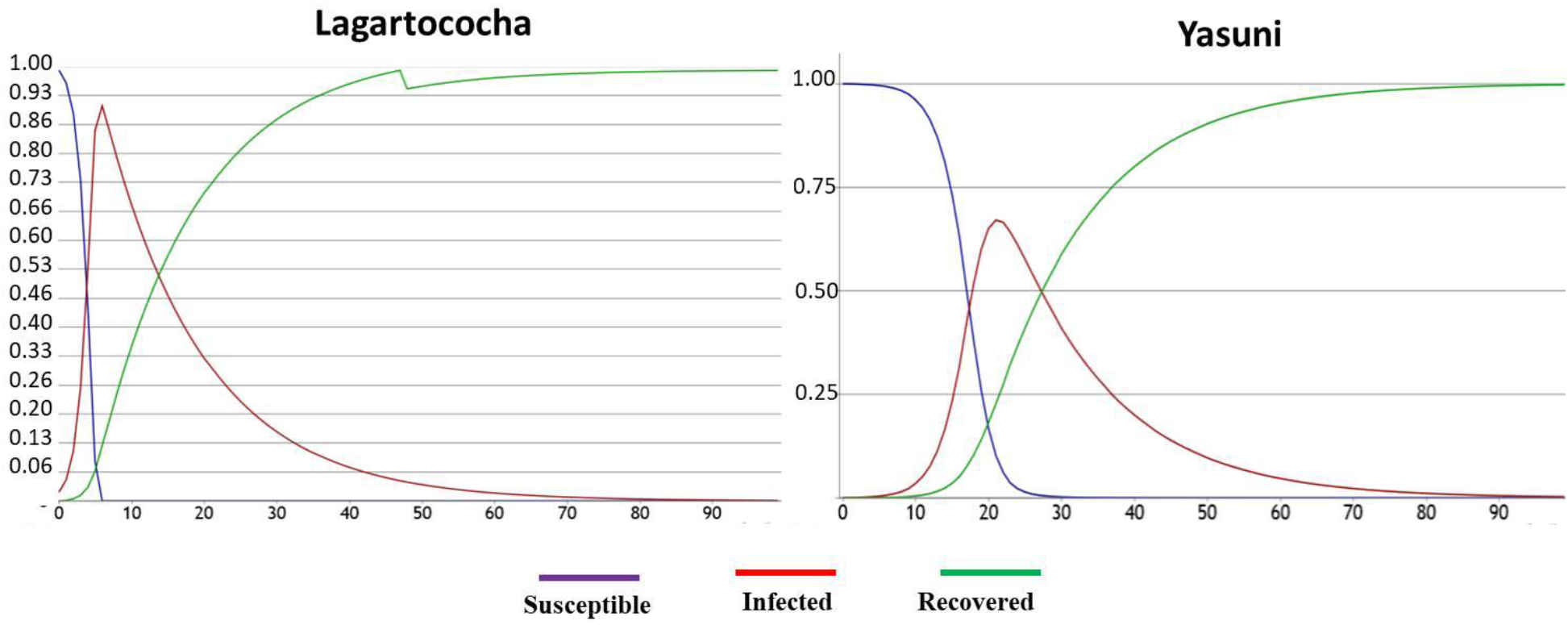
The COVID 19 SIR model of indigenous tribe populations living in remote Lagartococha and Yasuni rainforest enclaves (x axis: days, y axis: proportion of total population).

The simulated SIR models to show the flattened COVID 19 curve were also presented in Figure 4 for Lagartococha and Figure 5 for Yasuni populations. The simulations were made based on the assumptions if the interventions were applied. The model used 25%, 50%, and 75% intervention scenarios. As the model notices, an increase in the interventions is associated with a decrease in the infected cases and a flattening in the COVID 19 curve. Besides, the model shows a decrease in the proportion of total population infected by the COVID 19 and the disease lasts for a shorter period. For the Lagartococha populations, the effect is clear for 50% and 75% interventions. The peak of infected case is reduced from 86% to 73% under 50% interventions. Likewise, cases are reduced to 53% under 75% intervention scenario or almost half of the infected cases if there is no intervention applied.

**Figure 4.**
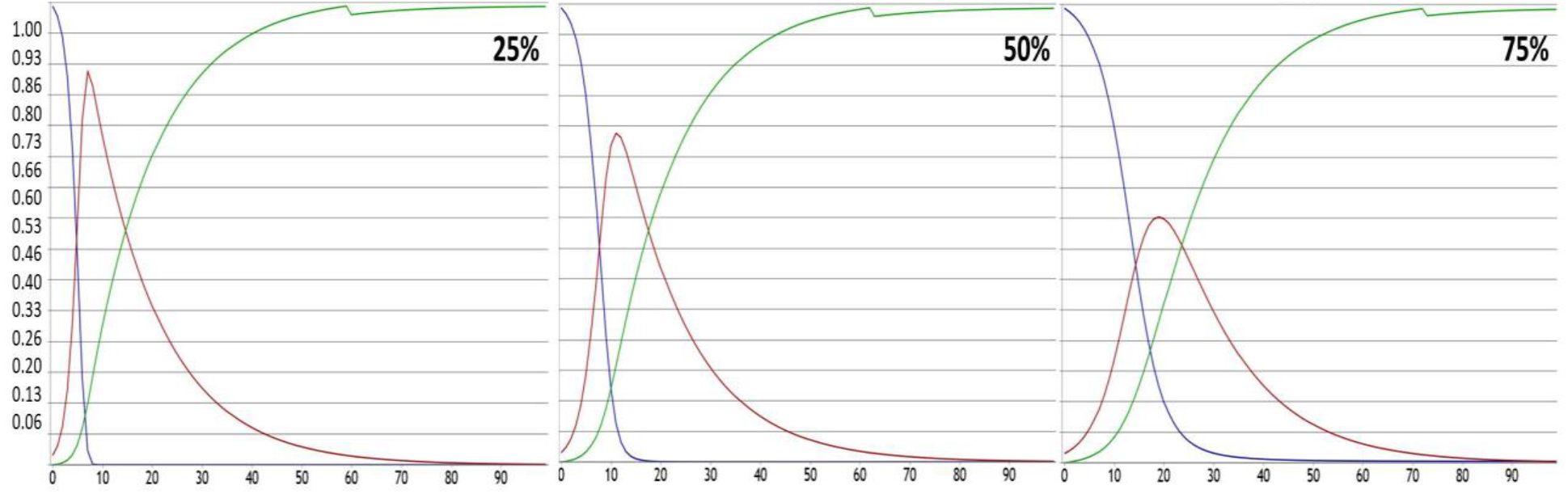
The COVID 19 SIR model of indigenous tribe populations living in remote Lagartococha rainforest enclaves with simulated 25% (low), 50% (moderate), and 75% (high) interventions (x axis: days, y axis: proportion of total population).

A flattened curve is observed for the Yasuni population as well (Figure 5). For the 25% intervention scenario, the COVID 19 peak is already reduced from 66% to 56%. The reductions are keep increasing for 50% and 75% interventions as well. For 50% intervention scenario, the proportion of infected cases are only 28% of the total population. While, COVID 19 peak is reduced to only 15% under 75% intervention scenario.

**Figure 5.**
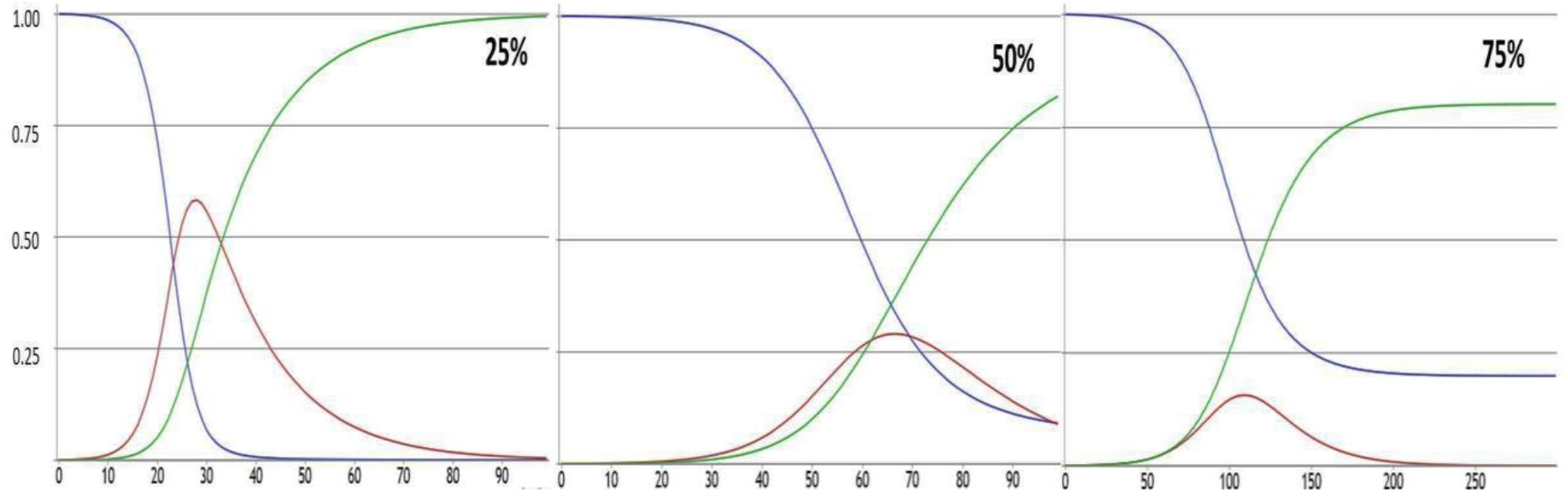
The COVID 19 SIR model of indigenous tribe populations living in remote Yasuni rainforest enclaves with simulated 25% (low), 50% (moderate), and 75% (high) interventions (x axis: days, y axis: proportion of total population).

## 4. Discussions

An indigenous tribe is accounted for significant populations living in the Amazon rainforest. It was estimated that there were 8.5 million indigenous people lived in Amazon (Hern 1991). Among them, 300000 people under 100 tribes are living in isolated parts of Amazon (Baumgartner *et al*. 2020). The population numbers are varied and dynamic. In this study, the indigenous populations are varied ranging from 744 to 4000. The proposed SIR model in this study simulates the widespread of COVID throughout indigenous tribe populations living in remote Lagartococha and Yasuni rainforests. The study gains insights from current data driven approach on epidemics in infected populations. Further this study recommends a measure to control the epidemic and to derive solutions for planning and managing confirmed cases.

The Figure 2 shows the SIR model applied to forecast the COVID 19 cases in Lagartococha and Yasuni rainforest enclaves based on the data available. From Figure 2, it is clear that the COVID 19 cases will grow rapidly over time resulting a vastly infected population and the situation may even get out of control if no necessary steps are taken. The COVID 19 cases in this study are comparable from other findings. It is estimated that the proportion of COVID 19 cases among indigenous populations is approximately 0.79% (Ortiz-Prado *et al*. 2020) and even 8.5% (Baumgartner *et al*. 2020). Therefore, it is high time to take precautions to reduce the COVID 19 and flatten the epidemic curve.

The SIR model shows that the peak of epidemic curve in Lagartococha tribe could possibly happens immediately in 10 days (Figure 3). While the peak for Yasuni population will occur after 20 days. Nonetheless, the result and the pandemic curve possibly contains the essential uncertainty due to the possibility of changes in the social and climatic situations. The peak of epidemic curve can be flattened by improving policies like travel restriction, sanitization, social distancing, and more testing measures. Flattening the curve refers to policy and behavior to control the COVID 19 daily cases at a manageable thresholds for health services. Flat curve assumes the numbers of COVID 19 cases but the cases are occurring slower over a longer period of time. A slower infection rate will reduce the stress of health services and hospital visits on any given day.

A SIR model developed by Wagh *et al*. (2020) has informed how the authority intervention can flatten the curve of COVID 19 in India. In the intact condition, the COVID 19 cases were infecting 30% of population within 60 days. Nonetheless, the SIR model with the authority intervention shows the COVID 19 cases were reduced to 15% and this happens within 90 days. Likewise in Bangladesh, Islam *et al*. (2020) have showed the case reductions due to the interventions. The lockdown interventions are estimated can reduce the cases from 20% to 50%. It reduces the infected population drastically which means social distancing is a major key to face this epidemic in this scenario. A similar flattened curve pattern was also observed in Cameroon, Egypt, and Malaysia (Arifin *et al*. 2020, Mahmud and Lim 2020). More than half reductions of the contacts with the infected individuals are simulated and proven can reduce the proportion of cases drastically from 90% to 30% in Cameroon (Nguemdjo *et al*. 2020). In Egypt (Hasab 2020), the interventions have reduced the cases from 28% to 1%. There are several public health interventions that can be adopted to the model and used to flatten the COVID 19 curve. Those simulated interventions are increasing social distancing through reductions of numbers of exposure to potential infected individuals on daily basis. The exposure numbers can be articulated as high exposure (25% with low interventions) and 75% with high interventions (low exposures). Moreover, the second simulated interventions are increasing hygiene measures through increasing frequency of hand washing like washing frequently (75% interventions) or never washing hands (25% interventions) (Tartari *et al*. 2019).

A confirmed COVID 19 especially in indigenous population (Figure 2) should raise a concern considering its isolation, population size, and genetic diversity. A study by Cardoso *et al*. (2012) confirmed the extremely low genetic diversity and low population size observed in the tribe population living in Yasuni forest. This condition related to the genetic drift events promoted by founder effects and anthropogenic factors includes tribe’s warlike customs. The same condition is also observed in the Callarú population. There was a decrease of the internal intervillage genetic heterogeneity in the Callarú as reported by Neel *et al*. 1980. This condition was caused by the migrations from the interior forest enclaves to the river bank and individual mobility (Salzano *et al*. 1980).

Under the COVID 19 pandemic threats, a population with the low genetic diversity may be more susceptible. Mulligan *et al*. (2004) found that low gene pools in native American tribes were determinant factors for common complex diseases, diabetes, and rare Mendelian disorders as well. The COVID 19 is a respiratory and systemic illness (Lippi and Henry 2020) and there was a history of emerging respiratory tract infections (Butler *et al*. 2001) and introduced diseases including tuberculosis (Hern 1991) among indigenous populations. In fact, the influenza and cardiorespiratory were common among Amazon tribes. As reported by Walker *et al*. (2015), influenza was accounted for 25% of observed disease in indigenous population. While Herndon *et al*. (2009) found that cardiorespiratory related diseases was accounted for 15.9%. A historical record of respiratory illness among indigenous tribes is related to the presence of ACE2 receptors. This is a concerning issue since the ACE2 is the receptor for COVID 19 (Zhang *et al*. 2020). Hakim and de Soto (2020) have identified elevated levels of ACE2 receptors in the lungs among indigenous peoples.

The individual health status is not only the determinant factors that can make the tribe indigenous populations become susceptible to the pandemic or can cause a burden to the prevention and mitigation efforts. There are several external factors that can contribute more to the spread of the diseases. Those factors include the availability of remote health centers and the rapid encroachment development. According to the INEI Indigenous Census in 2017, only 5% of indigenous populations in the entire Amazon have access to health facilities without hospitalization. Likewise only 1.7% have inpatient health centers. In regards to the rapid developments, according to Finer *et al*. 2009, currently there are several roads that can access the Yasuni remote populations. This road can increase the contacts with the individuals outside the intact indigenous populations and make this populations more prone to the disease.

## 5. Conclusions

This present study is the first that delivers the analyses of the COVID 19 dynamic focusing on the tribe populations using the SIR model. In here, the model has been presented by using several SIR input parameters. The model shows an intact exponential increase in the total number of infected cases. The COVID 19 cases will peak within 10 days for Lagartococha and 20 days for Yasuni populations. This gives a first idea of how the epidemic is spreading rapidly across the indigenous populations. By applying intervention scenarios from the lowest to the highest intervention levels to the SIR model, the epidemic peak can be reduced and hence curve flattens. This will indeed flatten the curve but will prolong the duration of the epidemic. With the simulated interventions, the COVID 19 peak can be reduced to more than half of the numbers of cases without interventions. These findings underline the importance of appropriate interventions for limiting and even stoppig the spread of the COVID 19 especially among the vulnerable and isolated indigenous populations.

## 6. Recommendations

This study provides room for improvements for estimating the effectiveness of health interventions. The administration combined with controlling authority intervention will help to flatten COVID 19 epidemic curve. As per the situation, intervention by populations supported with government policy for instance social distancing, it can be used to predict the reduction of final epidemic size using SIR model. The SIR model that used in this study can be easily applied to model the COVID 19 in other indigenous tribe populations for example in Africa, Australia, and Asia continents.

## Data Availability

Data used in this study are available in the manuscript

## Notes

### Competing Interest Statement

The authors have declared no competing interest.

### Author Declarations

Univ. of Indonesia

## References

Arifin WN, Chan W, Amaran S, Musa K. 2020. A Susceptible-Infected-Removed (SIR) model of COVID-19 epidemic trend in Malaysia under Movement Control Order (MCO) using a data fitting approach. *medRxiv*. 2020.05.01.20084384.

Baumgartner MT, Lansac-Toha FM, Coelho MTP, Dobrovolski R, Diniz-Filho JAF. 2020. Social distancing and movement constraint as the most likely factors for COVID-19 outbreak control in Brazil. *medRxiv*. 2020.05.02.20088013.

Belfin RV, Bródka P, Radhakrishnan BL, Rejula V. 2020. COVID-19 peak estimation and effect of nationwide lockdown in India. *medRxiv*. 2020.05.09.20095919.

Butler JC, Crengle S, Cheek Leach AJ, Lennon D, O’Brien KL, Santosham M. 2001. Emerging Infectious Diseases Among Indigenous Peoples. Emerging Infectious Diseases. 7(7):554–555.

Cardoso S, Alfonso-Sánchez MA, Valverde L, Sánchez D, Zarrabeitia MT, Odriozola A, Martínez-Jarreta, B, de Pancorbo MM. 2012. Genetic uniqueness of the Waorani tribe from the Ecuadorian Amazon. Heredity. 108(6):609–615.

Crosby AW. 1976. Virgin soil epidemics as a factor in the aboriginal depopulation in America. William Mary Q. 33: 289–299.

Finer M, Vijay V, Ponce F, Jenkins CN, Kahn TR. 2009. Ecuador’s Yasuní Biosphere Reserve: a brief modern history and conservation challenges. Environ. Res. Lett. 4

Hakim ST, de Soto JA. 2020. Medical Basis for Increased Susceptibility of COVID-19 among the Navajo and other Indigenous Tribes. Preprints. 202004.0217.v1.

Hasab AA. 2020. Flattening COVID-19 Curve in Egypt: An Epidemiological Modelling. Preprints. 202005.0156.v2.

Hern WM. 1991. Health and demography of native Amazonians: historical perspective and current status. Cad. Saúde Pública. 7(4): 451–480.

Herndon CN, Uiterlo M, Uremaru A, Plotkin M J, Emanuels-Smith G, Jitan J. 2009. Disease concepts and treatment by tribal healers of an Amazonian forest culture. Journal of ethnobiology and ethnomedicine: 5: 27.

Islam, Md, Ira J, Kabir K, Kamrujjaman Md. 2020. COVID-19 Epidemic Compartments Model and Bangladesh. *Preprints*. 202004.0193.v1.

Jo H, Son H, Hwanga HJ, Jung SY. 2020. Analysis of COVID-19 spread in South Korea using the SIR model with time-dependent parameters and deep learning. *medRxiv*. 2020.04.13.20063412.

Joralemon D. 1982. New World depopulation and the case of disease. J. Anthropol. Res. 38: 108–127.

King K, Lively C. 2012. Does genetic diversity limit disease spread in natural host populations?. Heredity. 109: 199–203.

Knibbs LD, Sly PD. 2014. Indigenous health and environmental risk factors: an Australian problem with global analogues?. Glob Health Action. 7.

Liddell C, Barrett L, Bydawell M. 2005. Indigenous representations of illness and AIDS in Sub-Saharan Africa. Soc. Sci. Med.60(4):691–700.

Lippi G, Henry BM. 2020. Chronic obstructive pulmonary disease is associated with severe coronavirus disease 2019 (COVID-19). Respiratory Medicine. 167.

Mahmud A, Lim P. 2020. Applying the SEIR Model in Forecasting The COVID-19 Trend in Malaysia: A Preliminary Study. *medRxiv*. 2020.04.14.20065607.

Mulligan CJ, Hunley K, Cole S, Long JC. 2004. Population genetics, history, and health patterns in native Americans. Annu. Rev. Genomics. Hum. Genet. 5: 295–315.

Nguemdjo UK, Meno F, Dongfack A, Ventelou B. 2020. Simulating the progression of the COVID-19 disease in Cameroon using SIR models. *medRxiv*. 2020.05.18.2010555.

Neel J, Gershowitz H, Mohrenweiser H, Amos B, Kostyu D, Salzan F, Smouse P, Oliver WJ, Spielman R, Mestriner MA, Lawrence D, simoes AS. 1980. Genetic studies on the Ticuna, an enigmatic tribe of Central Amazonas. Ann. Hum. Genet. Lond. 44: 37.

Ohenjo N, Willis R, Jackson D, Nettleton C, Good K, Mugarura B. 2006. Health of Indigenous people in Africa. Lancet. 367: 1937–1946.

Ortiz-Prado E, Simbana-Rivera K, Diaz AM, Barreto A, Moyano C, Arcos V, Vasconez-Gonzalez E, Paz C, Simbana-Guaycha F, Molestina-Luzuriaga M, Fernandez-Naranjo R, Feijoo J, Henriquez AR, Adana L, Lopez-Cortes Sr. A, Fletcher I, Lowe R, Gomez-Barreno L. 2020. Epidemiological, socio-demographic and clinical features of the early phase of the COVID-19 epidemic in Ecuador. *medRxiv*. 2020.05.08.20095943.

Salzano FM, Callegari Jacques SM, Neel JV. 1980. Genetic demography of the amazonian Ticuna Indians, Journal of Human Evolution. 9 (3).

Tartari E, Fankhauser C, Peters A, Sithole B, Timurkaynak F, Masson-Roy S, Allegranzi B, Pires D, Pittet D. 2019. Scenario-based simulation training for the WHO hand hygiene self-assessment framework. Antimicrobial Resistance & Infection Control. 8(1).

Vos T, Barker B, Begg S, Stanley L, Lopez AD. 2009. Burden of disease and injury in Aboriginal and Torres Strait Islander Peoples: the Indigenous health gap. Int. J. Epidemiol. 38(2):470–477.

Walker RS, Sattenspiel L, Hill KR. 2015. Mortality from contact-related epidemics among indigenous populations in Greater Amazonia. Scientific reports 5: 14032.

Wagh CS, Mahalle PN, Wagh SJ. 2020. Epidemic Peak for COVID-19 in India, 2020. *Preprints* 2020050176.

Waqas M, Farooq M, Ahmad R, Ahmad A. 2020. Analysis and Prediction of COVID-19 Pandemic in Pakistan using Time-dependent SIR Model. *arXiv*. 2005.02353v1.

Zhang H, Penninger JM, Li Y. 2020. Angiotensin-converting enzyme 2 (ACE2) as a SARS-CoV-2 receptor: molecular mechanisms and potential therapeutic target. Intensive Care Med. 46: 586–590.

Zhao Z, Li X, Liu f, Zhu G, Ma C, Wang L. 2020. Prediction of the COVID-19 spread in African countries and implications for prevention and control: A case study in South Africa, Egypt, Algeria, Nigeria, Senegal and Kenya. Science of The Total Environment. 729(10).

